# *‘When will it ever be enough?’* Black mothers’ and community advocates’ perspectives on why evidence of racism in English maternity care does not lead to action: a co-produced focus group study

**DOI:** 10.64898/2026.11.13.26368002

**Authors:** Sarindi Aryasinghe, Susan Ibuanokpe, Rhianna Newby-Mayers, Phayza Fudlalla, Carole Waithe, Gabriella Sarpong, Ruchi Wadhwa, Phoebe Averill, Sabrina Das, Helen Ward

## Abstract

**Background:** Racism shapes the maternity care and outcomes of Black, African, Caribbean and mixed-Black women in the UK. Services monitor inequities through outcomes and experience data, yet evidence of racism often results in inaction, and existing explanations rely mainly on staff perspectives. We drew on Black mothers’ and community advocates’ accounts to explain what stops monitoring of racism from leading to action, and how they propose evidence of racism should be recorded so that it does.

**Methods:** This co-produced qualitative study was underpinned by critical realism and Black feminist thought. Four focus groups were held in London from June to July 2026 with nine Black, African, Caribbean and mixed-Black mothers who used NHS maternity services and seven community advocates. Lived experience co-researchers co-led the study design, data collection and analysis. We used critical realist thematic analysis to propose mechanisms explaining participants’ accounts, each tested against rival explanations.

**Results:** We propose three mechanisms: credibility discounting, evidence neutralisation and containment of change. Participants described Black women’s accounts being discounted until someone else raised the same concern, consistent with testimonial injustice. Recorded evidence appeared to be neutralised through audit culture, as concerns were acknowledged or deflected without action. Change that would shift power or accountability appeared to be contained, consistent with institutional racism. Participants also described harm from staff who shared their racial identity, linked to internalised racism intersecting with language and migration, and to workplace cultures in which Black staff are also dismissed; we interpret this harm as a possible outcome of these mechanisms. Participants proposed that services account for their response to each concern, supported by community-generated evidence, independent observation, qualitative routes and ethnicity-disaggregated data. This would require protection for those who raise concerns and sustained funding and backing for action.

**Conclusions:** Our findings suggest more evidence alone is unlikely to address racism in maternity services. An independently reviewed record of how services respond to each concern could show when they do not act. Change is more likely where services shift power and resources to community organisations working with Black families, while remaining accountable for acting on evidence of racism.

## 1. BACKGROUND

Racism shapes the maternity care and outcomes of Black, African, Caribbean and mixed-Black women in countries where they are racially minoritised [1–3]. In the UK, Black women are up to three times more likely than white women to die during pregnancy or up to six weeks after birth [3]. Racism operates as a system that distributes opportunity and worth according to how people are racialised [4]. In maternity care, this system is visible in Black women’s accounts of their concerns being dismissed, ignored or disbelieved [5–7]. A national investigation across twelve English National Health Service (NHS) organisations found racism ingrained throughout maternity services [8].

Much of the policy response relies on monitoring, by which we mean the routine ways services collect evidence of racism. These include complaints, patient experience surveys and the NHS Maternity and Neonatal Equalities Dashboard, which reports outcomes and experience by ethnicity and deprivation [9]. These approaches can show where care differs between groups or record individual concerns, but they say little about the decisions behind those differences or how services should improve. England’s national maternity and neonatal investigation by Baroness Amos identified racism as a ‘critical maternity safety issue’, recommending that inequalities data be captured as safety intelligence and escalated to board level [8]. This approach assumes that recorded evidence prompts action. However, one NHS organisation was aware of poorer experiences and outcomes for racially minoritised women for over a decade, while initiatives to address them were inconsistently resourced [10].

Existing research partly explains why evidence goes unused. Studies of patient feedback and speaking up in the NHS suggest that staff are more likely to act on concerns when they value patient input and have the autonomy and resources to respond [11], and that staff may not raise their own concerns about safety or racism when they expect a defensive response and limited change [12,13]. Formal institutional commitments to anti-racism, such as policies and written values, can also take the place of action, as stating a commitment may be treated as equivalent to realising the change it promises [14]. Although patient-reported experience measures of obstetric racism and respectful maternity care have been developed [15,16], such evidence may do little if the barriers lie in how it is handled. Independent investigations have documented families’ concerns being dismissed [8,10,17], but research examining why concerns are not addressed has drawn largely on staff accounts.

These explanations have rarely been examined from the perspective of women and families who raise concerns about racism. Black mothers hold direct knowledge of how their concerns were responded to, while community advocates observe how concerns are handled across many families and services [18]. Co-production, in which researchers, practitioners and communities jointly conduct research [19], offers a way to bring this knowledge into decisions about how evidence of racism is collected and used. This study therefore asks, from the perspective of Black mothers and community advocates: (1) what appears to stop the monitoring of racism in maternity care from leading to action; and (2) how evidence of racism should be recorded so that it does.

## 2. METHODS

### 2.1. Study design and theoretical approach

We co-produced a qualitative focus group study underpinned by critical realism, which holds that reality exists independently of our knowledge of it and distinguishes between experiences, the events experienced and the causal mechanisms that produce them [20,21]. Because the social world is an open system, these mechanisms operate as tendencies rather than certainties [22], and our knowledge of these mechanisms is fallible and open to revision [20]. Social structures such as institutional racism are considered underlying mechanisms, which can be explored and inferred through qualitative methods [22,23].

Our study was also guided by Black feminist thought, which treats Black women’s lived experience as a criterion of meaning and holds that their experience of racism and sexism together gives them a particular view of how oppression operates. This knowledge is built and tested through dialogue with other Black women [24]. Critical realism complements this by explaining the mechanisms through which interlocking systems of oppression produce harm, including those that the people affected or institutions may not recognise [25]. This emphasis on dialogue informed our choice of focus groups. Groups who share identities can provide a more comfortable setting to discuss experiences [26], and experiences may only be recognised as racism when discussed with others who have had similar ones [27]. Group interaction also allows participants to build on each other’s accounts [28]. Black feminist thought also informed our co-produced approach, in which five lived experience co-researchers from Black, African, Caribbean and mixed-Black backgrounds contributed to the study design, topic guide, facilitation, coding and interpretation, as reported using the GRIPP2 short form (Additional file 1) [29].

However, combining the two creates tension. Black feminist thought treats the knowledge of people whose voices are rarely heard as authoritative, whereas critical realism holds that racism exists whether or not it is recognised, so researchers may need to look beyond participants’ own explanations [25,30,31]. We addressed this by sharing responsibility for exploring mechanisms and rival theories with the lived experience co-researchers during coding and interpretation. Key findings were shared with participants for review, which further refined our interpretations.

Study reporting adheres to the Standards for Reporting Qualitative Research (Additional file 2) [32].

### 2.2. Our positionality and reflexivity

SA, a South Asian woman and doctoral researcher, works for a national anti-racism organisation in health. This gave insight into how services use evidence of racism, but her organisation’s commitment to anti-racism risked leading her to read participants’ accounts as evidence of racism before other explanations had been considered. SA also does not share participants’ experiences of anti-Black racism or of NHS maternity care. SA therefore kept reflexive memos and developed interpretations with the five lived experience co-researchers, who had used NHS maternity care or supported Black families as advocates and had worked with SA for two years. Their own experiences, discussed in fortnightly meetings, informed findings only where participants’ accounts supported them. RW, a South Asian woman and patient and public involvement professional, supported recruitment and coding. SA’s supervisors reviewed and challenged interpretations; HW is a white British woman and clinical public health academic focused on improving research and healthcare for marginalised populations, PA a white British woman and researcher in healthcare quality and safety, and SD a mixed-race Southeast Asian woman, NHS obstetrician and reproductive health researcher. As the co-researchers were the only Black members of the team, their judgement was central to interpreting accounts of anti-Black racism.

### 2.3. Study setting

The study took place in London, where a higher proportion of residents are Black, African, Caribbean and mixed-Black than elsewhere in England [33]. Participants could therefore describe care across several maternity services, including those that have committed to addressing racism.

### 2.4. Ethics approval

The study received a favourable opinion from the Imperial College Research Ethics Committee (ref. 8208475). Participants gave signed electronic consent and could withdraw up to 14 days after their session, although contributions to a group discussion could not always be removed. Confidentiality was maintained unless a serious risk was disclosed, and participants were signposted to NHS complaints routes where relevant.

### 2.5. Participants and recruitment

We purposively sampled maternity service users aged 18 or over who had used London NHS maternity services within the past five years and self-identified as Black, African, Caribbean or mixed-Black. All participants identified as mothers, and so we refer to them as such. We also recruited community advocates of any ethnicity who currently support Black families in London during pregnancy, birth or the postnatal period. Participants were recruited through community and professional networks, social media and word of mouth. Eligibility was confirmed through an online screening questionnaire and a phone call with SA or RW, which also screened for fraudulent participation. Mothers and advocates took part in separate groups so that advocates’ knowledge of services would not shape how mothers described their own care.

We assessed sample adequacy using information power dimensions [34]. Our aim was narrow, concerning what happens to evidence of racism in maternity care in a defined population and setting, and our sample was specific to it. Established theories also guided retroduction towards specific mechanisms. Dialogue was rich, as lived experience co-researchers co-facilitated the groups and participants readily described their own experiences unprompted. Two of the groups were smaller, which allowed each participant more time to speak and may have created the safety needed to discuss experiences of racism more quickly [35]. Our cross-group analysis called for more data, but the narrow aim, specific sample and quality of dialogue offset this.

### 2.6. Data collection and processing

Four focus groups were held in June and July 2026, two with Black mothers and two with advocates. Each was facilitated by two lived experience co-researchers with SA and RW, all trained in trauma-informed facilitation and qualitative research, who debriefed after each session. The semi-structured topic guide, co-produced by SA and the lived experience co-researchers, asked about good and fair maternity care, what evidence could show racism, and how evidence could be captured and used; it was revised for clarity after the first two groups (Additional file 3). Participants received example questions and support resources in advance.

Participants could attend in person at a community venue in south London or online on Zoom (Table 3); in-person groups were recorded with a dictaphone. Sessions lasted 90 to 120 minutes and were conducted in English. Ground rules were agreed, and questions allowed participants to describe scenarios rather than their own experiences of racism, although many shared their own. Participants received a £50 thank-you voucher.

Recordings were pseudonymised, stored on encrypted servers and professionally transcribed after a 14-day withdrawal period. SA checked transcripts against the recordings and removed identifying details before analysis.

### 2.7. Coding and data analysis

We used critical realist thematic analysis as described by Fryer [21], which aims to produce causal explanations of events rather than descriptions of experience and draws on existing theory to assess and strengthen explanations derived from the data [36]. Following this distinction, participants’ accounts record their experiences, codes consolidate these into events (what happened), and themes explain the mechanisms through which these events occur [21]. We followed Fryer’s five steps (Table 1), moving between them iteratively.

**Table 1.** Application of Fryer’s five-step approach to our critical realist thematic analysis [21].

|  | Explanation | What we did |
|---|---|---|
| **Step 1: Develop your research questions** | Identify the experiences and/or events of interest and develop one or more research questions on causal mechanisms | Our first research question asks what appears to stop the monitoring of racism in maternity care from leading to action; our research questions were refined and narrowed during analysis, which is common in qualitative thematic analysis [43]. |
| **Step 2: Data familiarisation** | Read the transcripts, record initial thoughts and questions, and note details around data collection | SA, RW and the lived experience co-researchers (SI, CW, GS, RNM and PF) read and listened to all transcripts and recordings, taking notes in a reflexivity journal on initial thoughts, questions, contradictions, and 'silences' in the data. |
| **Step 3: Apply, develop, and review codes** | • Apply descriptive codes that capture experiences and events as described by participants, and any mechanisms offered by participants as explanations.<br>• Review and consolidate codes iteratively. | SA, RW and the lived experience co-researchers (SI, CW, GS, RNM and PF) reviewed excerpts from all transcripts together in a series of coding workshops and fortnightly meetings, discussing and applying codes as a group using Microsoft Word.<br>SA then consolidated the codes, which were reviewed by RW and the lived experience co-researchers in subsequent fortnightly meetings, and applied them to the full transcripts in NVivo v.15. |
| **Step 4: Develop and review themes** | Develop themes that demonstrate potential causal mechanisms, and review their theoretical validity (are the claims logical, and do they appropriately explain what is in the data) [41]. | Drawing on discussions with all co-authors, the explanations offered by participants and established theory, SA developed candidate causal mechanisms and tested each against rival explanations and cases in which it did not operate. These were refined through feedback from all co-authors and findings review sessions with interested focus group participants, where SA, SI, PF and CW presented the findings. |
| **Step 5: Generate conclusions and report** | Consider the analysis' overall contributions and implications, and approaches to communicate the findings | In addition to an academic publication, findings are shared in summarised form for maternity policymakers and communities. |

Codes were consolidated through abduction, reinterpreting the events participants described through theoretical concepts from the literature [22,36]. Themes were developed through retroduction, working back from those events to the structures and mechanisms that could have produced them [37,38]. We organised the mechanisms using Jones’s levels of racism [39], with other theories to explain how racism operates at each level. For each consolidated event, we asked Fryer’s question, ‘what best explains this event?’ [21], and under what conditions the proposed mechanism takes effect [36]. As the topic guide asked participants why evidence was not being used, their own causal explanations were the starting point.

These were scrutinised against the events described and extended where theory or other accounts supported a fuller explanation [30,36]. Each mechanism was tested against rival explanations [21,40]. Because mechanisms operate as tendencies [22,37], all transcripts were searched for cases in which the expected outcome did not occur.

During consolidation, codes were reviewed for interpretative validity, checking that the broader concepts still reflected participants’ experiences [21,41]. Themes were reviewed for theoretical validity [41], meaning whether each causal explanation rested on sound reasoning (judgemental rationality) and explained the patterns found across the data (explanatory power) [40]. Rigour therefore came from reflexive attention to validity rather than from agreement between independent coders [21,40]. Joint coding was used to test interpretations against lived experience.

As interpretation is integral to critical realist thematic analysis, we present the proposed mechanisms, rival explanations and their theoretical grounding within the Results [42]; the Discussion therefore focuses on how they extend existing research and their implications.

## 3. RESULTS

### 3.1. Participant characteristics

Sixteen participants, nine Black mothers and seven advocates (Table 2), took part in groups of two to six. Two groups were smaller than planned because of drop-outs on the day.

**Table 2.** Focus group participant characteristics.

| Characteristic |  | Black mothers (n = 9) | Advocates (n = 7) |
|---|---|---|---|
| Ethnicity (self-described) | Black African | 7 | 1 |
|  | Black British African | 1 | 0 |
|  | Black Caribbean | 0 | 3 |
|  | Mixed Black African and Caribbean | 1 | 2 |
|  | Asian | - | 1 |
| Gender (self-described) | Woman | 9 | 6 |
|  | Man | 0 | 1 |
| Age | 18 to 29 years | 3 | 0 |
|  | 30 to 39 years | 4 | 3 |
|  | 40 and over | 1 | 4 |
|  | Prefer not to say | 1 | 0 |
| Time in the UK | Less than 2 years | 2 | 0 |
|  | 2-10 years | 2 | 0 |
|  | More than 10 years | 0 | 1 |
|  | Born in the UK | 4 | 6 |
|  | Prefer not to say | 1 | 0 |
| Number of pregnancies NHS maternity services were used† | One | 5 | 1 |
|  | Two | 3 | 2 |
|  | Three or more | 1 | 2 |
|  | Prefer not to say | 0 | 1 |
|  | Not applicable | 0 | 1 |
| Time as an advocate | Less than 4 years | - | 3 |
|  | 4 years or more | - | 3 |
|  | Prefer not to say | - | 1 |
| Type of advocacy role* | Charity, CIC, community group | - | 5 |
|  | NHS maternity service user representative role | - | 3 |
|  | Independent or freelance | - | 2 |
|  | Local authority-affiliated | - | 2 |

*\*The numbers do not add up to n=7 as an advocate can have more than one role.*
*†Advocates answered about their own pregnancies, as some had also used NHS maternity services*
*CIC, community interest company; NHS, National Health Service.*

Quotations are labelled by participant identifier and role (Table 3).

**Table 3.** Focus groups and participant IDs.

| **Focus group** | **No. of participants** | **Participant IDs** | **Mode** |
|---|---|---|---|
| Group 1 (Mothers) | 6 | P1-P6 | In-person |
| Group 2 (Mothers) | 3 | P7-P9 | Online (Zoom) |
| Group 3 (Advocates) | 5 | P10-P14 | Online (Zoom) |
| Group 4 (Advocates) | 2 | P15-P16 | In-person |

### 3.2. What stops current monitoring of racism in maternity care from leading to action

We identified three mechanisms that may limit the extent to which monitoring of racism in maternity care leads to action, each acting at a different point between a concern being raised and services changing (Figure 1). Black women’s accounts are discounted, so their concerns are often not recorded; what is recorded is neutralised through NHS audit processes; and change that would shift power or accountability is contained. All three mechanisms were described in all focus groups. Participants in three groups also described harm from staff who share a racial identity with the women they care for, which we interpret as an outcome of the same mechanisms.

**Figure 1.**
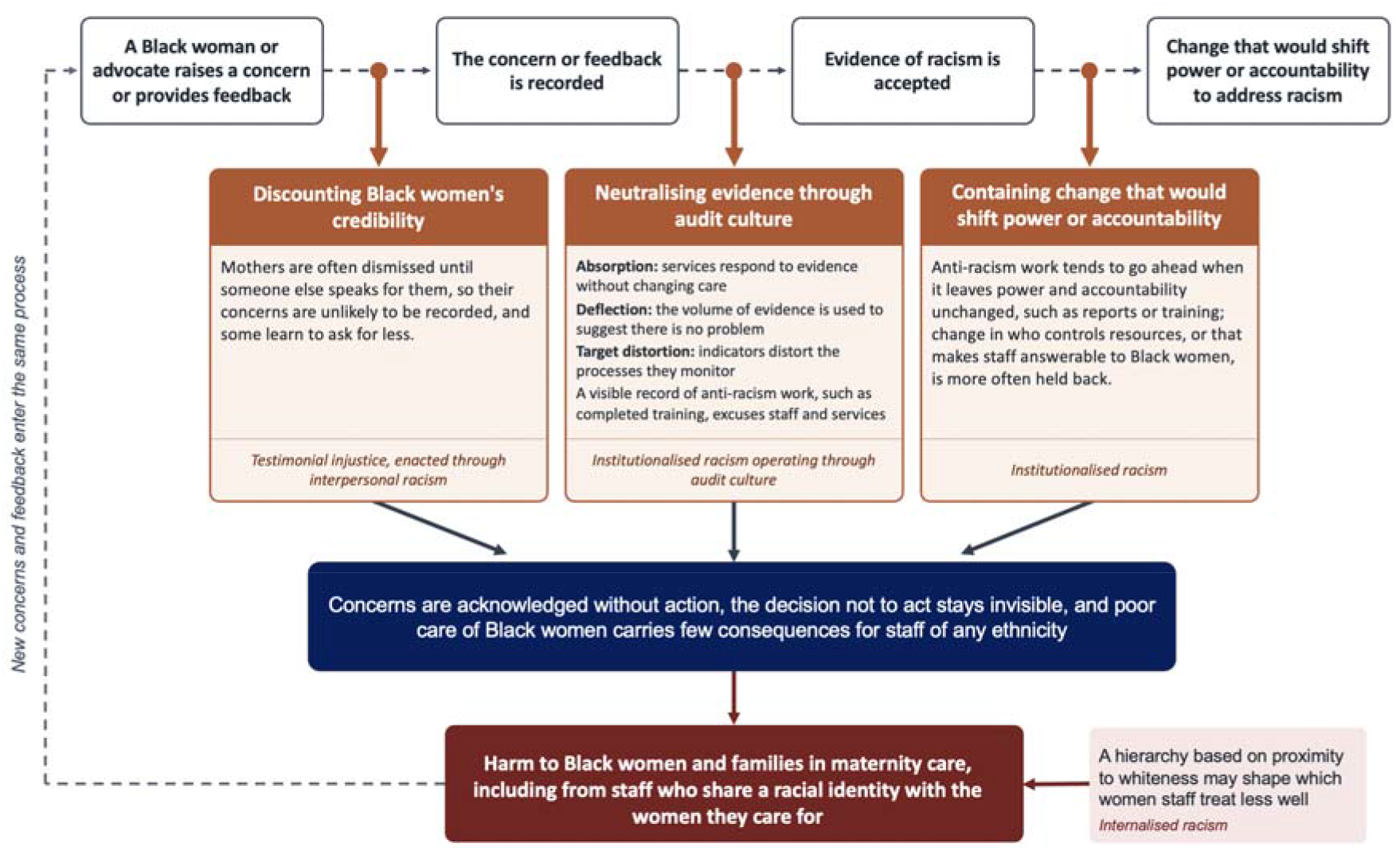
Proposed mechanisms through which evidence of racism in maternity care does not lead to action

#### 3.2.1. Discounting Black women’s credibility

Participants described how the same concern could receive a different response depending on who raised it, with mothers often dismissed until someone else spoke for them. One mother was left in pain during her induction until a visitor to another woman fetched help. She believed that without a student who asked to examine her once more, she would have been taken for an emergency caesarean section: ‘If not for that student, I am sure they would have cut me open’ (P1, mother). Another mother, who laboured for almost twenty-four hours with only oral painkillers, linked this to the racial stereotype of Black women having higher pain thresholds.

Over time, women described adapting how they sought care. One mother recalled a receptionist who was ‘so friendly and so lovely’ to white women waiting but obstructive towards her, and who changed her manner once a white obstetrician intervened. On a later visit, she ‘felt like I had to really over-explain why I’m here’ (P7, mother). Others brought someone with them to be heard:

> *‘When we go to the hospital… we have to come with somebody, normally a male figure, to be listened to, to be respected. Because us on our own, it’s like we’re not seen as… just oftentimes, not taken seriously.’* – P9 (mother)

In the same focus group, mothers debated whether self-advocacy should be their burden to carry:

> P7: ‘…the *onus is on the mother as well to understand and be able to, kind of, express themselves in a way that I guess those who are looking after them can interpret, and understand, and receive.’*
>
> P9: ‘…I *agree with that, but then where does that information come from? It comes from the hospital… even in that information being shared, sometimes they do gatekeep information as well.’*
>
> P7: ‘100%, *I agree with you, but I think that’s just how the NHS system is designed…*’
>
> P8: ‘I *feel like even how we’re talking now, it’s almost like we’re always preparing for a battle… we have to because it can be life or death… if you’re not someone that can advocate for yourself then you might end up getting the worst care ever because you just take what you’re given…*’

In this exchange, self-advocacy shifted from an individual responsibility to a response to services that withhold information and do not listen. It is therefore an unreliable safeguard for women least able to advocate for themselves. Later, one mother suggested that being dismissed teaches women ‘to ask for less so that they don’t feel like a burden’ (P9, mother). Another described holding back a complaint because ‘it’s going to be one word against the other’ (P7, mother).

One advocate described Black fathers who spoke on behalf of their partners being seen as aggressive and removed from the ward rather than heard. An advocate also described a woman awaiting rehousing who said she felt too unwell to be discharged and was told that staying would not get her housing; she was later found to have a major infection (P15, advocate). Where harm occurred, the burden of being heard could also be turned back on parents. One advocate witnessed staff at times blaming parents after babies had died: ‘They would say, “She didn’t say she was in pain. She didn’t say it loud enough. She didn’t come in enough times when her baby stopped moving.”’ (P13, advocate). In one example, a concern that services had ignored was acted on quickly once an advocate raised it: ‘the same admin team who hadn’t replied for weeks, then got her an appointment straight away’ (P16, advocate). Conversely, one mother described a midwife in a later pregnancy who knew her history and, without her knowing, called the ward to ensure staff looked after her (P2, mother). Continuity of carer may help concerns to be heard, although here too a professional spoke for the woman.

We tested three rival explanations. Professional hierarchy could explain some dismissals, as most of those dismissed were service users speaking to clinicians, and staff concern about the extra work a request might create could explain others. Neither explains why a receptionist with no clinical authority treated white and Black women differently while they waited for the same service, or why Black fathers advocating for their partners were seen as aggressive rather than worried. A third, clinical judgement, does not explain why the woman with a major infection was dismissed. We therefore interpret this mechanism as testimonial injustice, in which a speaker is given less credibility because of prejudice about their social group [45], enacted through interpersonal racism [39]. Participants’ accounts point to one source of this prejudice, images of Black women as strong, able to bear pain and aggressive, which Collins calls controlling images [24]. That a white obstetrician’s intervention changed how one mother was treated suggests whiteness acted as a credential, so that the obstetrician was listened to where the mother had not been [46]. Mothers who expect not to be believed may also narrow what they say [47], which may explain why women learned to ask for less. For monitoring, a concern that is not believed is unlikely to be recorded, and where harm occurs, responsibility may be placed on Black women themselves.

#### 3.2.2. ‘Sorry, and bye-bye’: neutralising evidence through audit culture

Where concerns were recorded, participants described three ways in which evidence was neutralised. In the first, which we term absorption, services responded to evidence without changing care. This resembles organisational decoupling, in which formal structures such as complaints processes confer legitimacy while remaining separate from practice [48], a process that is often racialised, allowing equity commitments to coexist with unchanged care [46]. After complaining, one mother received a written apology and a call from a lead midwife, with the harm attributed to a busy period. Another completed a feedback questionnaire, ‘but still yet they did it to somebody else’ (P4, mother). Advocates described women’s accounts reaching governance meetings without leading to change, leaving many with ‘a, “Sorry, and bye-bye,” and being expected to go into the world with all the trauma and all the pain’ (P12, advocate). Where advocates raised concerns, responses resolved the individual cases of those who attended service-led feedback meetings, leaving out ‘all the others who didn’t come’ (P16, advocate).

In the second, which we term deflection, services used low numbers of complaints or average figures to suggest there was no problem, pointing to their own data to show they were not outliers. Mothers noted that fewer complaints from Black families reflected long, difficult processes, with women ‘so fatigued… that they just can’t be arsed to report it’ (P8, mother). An advocate described how openness within a service did not reach board-level discussions:

> *‘…There is a gaslighting culture amongst even themselves. So I’ll go to an internal meeting and they’ll be more open on where something went wrong, and then you go to the board meeting, and they’re, “Oh, we did this, we did that.” So there is this culture of, “Yes, it went wrong, but we never really admit it.”’* – P16 (advocate)

The third, raised in both advocate groups, was target distortion, in which indicators used in decision-making distort the processes they monitor [49], so that ‘the numbers are good on your tables, but a lot of women are destroyed’ (P11, advocate). Two advocates, P15 and P16, developed an explanation together:

> *P16: ‘Because they’re just so scared…’*
>
> *P15: ‘Do you think if upper management financially lost out, that they’d be more…?’*
>
> *P16: ‘But they gain money from meeting targets that show that they’re, so-called, doing all of…’*
>
> *P15: ‘No, flipping it, inverting it. So for you to gain that money…’ P16: ‘You’d have to have evidence of certain outcomes.’*
>
> *P15: ‘And if the lived experience that’s coming through negates that, that’s a lot of evidence, like you’re saying.’*

Their exchange moved from leaders’ fear to incentives, which, they suggested, reward services for appearing to meet targets rather than for changing care. Participants also attributed this pattern to staffing shortages and to a defensive culture in which, ‘when they’re on the spot… they’ll never ever really say, “We messed up”’ (P16, advocate). As a rival explanation, defensive culture may explain why services avoid admitting fault more generally [8,10], but participants’ accounts suggest evidence of racism faces additional barriers. Complaints about racism often rested on one woman’s account against a professional’s. Two mothers described the evidence services required before acting:

> *P9: ‘…systemically they can make the evidence threshold so high or so unrealistic that even when you submit the evidence, it’s not enough for them to take accountability or to act.’*
>
> *P8: ‘…Like you said, the thresholds… When will it ever be enough? Which I think one concern is one concern too many if no one has listened to you… they just stack up all this data and they don’t actually do anything with it.’*

This suggests that collecting data may itself stand in for action. During analysis, the lived experience co-researchers proposed that staff may also be excused by a visible record of anti-racism work, such as completed training. We treat this as part of neutralisation, because diversity initiatives can serve a performative function while leaving the distribution of power unchanged [14]. Extending participants’ explanations, we interpret neutralisation as institutional racism operating through audit culture [39,50–52]. In audit culture, processes designed to demonstrate accountability turn evidence of racism into evidence of responsiveness, or set thresholds it cannot meet, while staffing pressures and fear of repercussions offer credible reasons why nothing could be done.

#### 3.2.3. Containing change that would shift power or accountability

Even where evidence of racism was accepted, participants described anti-racism work that left existing power and accountability unchanged, such as commissioning reports or cultural competency training, as more likely to go ahead. Change that would shift who controlled resources, or make staff answerable to Black women, was more often held back, most clearly where funding that could have gone to community organisations was kept within the NHS:

> *‘I’ve personally seen, in services before, where there is money… that could be given to Black and brown projects or the community as such, and instead what will happen is the NHS will decide to internally take on that budget… So, the money is there, but the allocation isn’t happening in the right way.’* – P14 (advocate)

Another advocate described a grant for Black maternal health in London that stalled after a disagreement between consultants and commissioners: ‘everybody who applied for the grant has just been left in hold, there’s no communication’ (P10, advocate). Participants described recommendations being held back in the same way; an advocate also noted that a recent independent review of a trust’s maternity services included a chapter on racism but no recommendation to tackle the issues raised [10].

Mothers offered possible explanations for this pattern. One located the decision not to act with senior leaders who ‘probably don’t look like any of us in this room… So, if it doesn’t affect them, or their community, or their people, why would they care?’ (P4, mother). Another suggested that current arrangements suited those who ran services, since ‘nobody is going to hold you accountable if you mistreat a Black woman’ (P9, mother).

Others attributed the lack of action to money, a rival explanation that participants themselves contested. One mother suggested that the problem ‘comes down to cost and money’ (P7, mother), but another questioned this, replying:

> *‘I disagree on the money thing, because some of these things don’t cost money. Manners, kindness, and respect are all free… I also think it’s because it benefits them. The way how things are, it doesn’t change their privilege, it doesn’t change their income, it doesn’t change nothing.’* – P9 (mother)

One mother argued that while services did not act, funding for equity work should be diverted to Black-led care, because ‘it’s not really fair to tell people to, “Oh, hang on. We’re just going to keep on with this racism a little bit longer,” when it’s a risk to life’ (P8, mother). She also suggested that inaction could be countered when ‘someone from your community in a high-up position’ took the issue on ‘as a moral thing to do’, although this would rely on individual commitment rather than accountability. An advocate also questioned why funding could be found during the COVID-19 pandemic but not for racism in maternity care. This comparison, together with accounts of funding being retained or left unspent and of no-cost changes such as respectful care not being made, suggests that the barrier lay in who controlled resources rather than whether they existed. We interpret this as institutional racism [39], in which routine processes maintain unequal access to power and resources without any individual needing to intend harm. As the mothers could not observe how funding decisions were made, this interpretation rests mainly on advocates’ accounts from inside services and on the pattern of what did and did not go ahead.

#### 3.2.4. Harm from staff who share a racial identity

As Figure 1 shows, the three mechanisms together may reproduce racism in maternity care, and when women raise concerns about the resulting harm, those concerns may be discounted, neutralised and contained in turn. Beyond well-documented harms [3,5,7], we focus on a less examined one, harm from Black staff, described in three groups. We interpret this harm as an outcome of the same mechanisms, which leave poor care of Black women with few consequences for staff of any ethnicity. In the first group, Black African mothers who had recently arrived in the UK described being treated worse by Black staff who had lived in the UK for longer:

> *‘…when I went for this sugar test, it was a Black woman that was acting as if she is racist… They believe that once they have stayed here for a long time, and they can speak like the whites, so they try to treat us that have just come, that we still speak, like, you know this person is from Africa, the way they treat us is very, very bad.’* – P6 (mother)

Others in the group agreed, although one mother, born in the UK, disagreed. Advocates explained the same behaviour through the system Black staff work in, with one describing how staff respond to the accountability they face:

> *‘When I come into the system, they’re telling me, “Hush, hush. Come on, you can manage,” because I look like their niece, I look like their sister, I look like their cousin… When a white woman comes in who they know will complain and who may give them problems, they treat her like she’s the princess of the world, and that is because they’re in a system that will put them into trouble if they don’t treat that white woman that way.’* – P12 (advocate)

Another advocate traced this treatment to a culture set from the top:

> *‘…When you speak to those midwives, because it’s a culture, that’s what’s coming from the top. Because when I’ve read the feedback of service users and I’ve read the feedback of some of those midwives who were leaving [their jobs], it’s exactly the same: “I was dismissed. I wasn’t listened to.”’* – P16 (advocate)

Mothers and advocates explained this harm in two ways, which we interpret as different levels of the same process. Mothers’ accounts suggest internalised racism, in which a hierarchy based on proximity to whiteness positioned women by their time in the UK and the way they spoke [39]. Here, racialisation intersected with migration [53]. Advocates’ accounts locate the harm in the system staff work in, where complaints carry consequences unequally and staff are dismissed as women are. This also reflects the national investigation’s finding that bullying, racism and discrimination by senior leaders and clinicians affect safety [8]. Internalised racism may influence which women staff treat less well; that the three mechanisms allow this treatment to continue rests mainly on advocates’ accounts.

### 3.3. Mothers’ and advocates’ proposals on evidencing racism in maternity care

Participants’ proposals focused on making visible what services decided, whose accounts were believed and what happened after a concern was raised, rather than on new measures of racism. We synthesised them into one framework (Figure 2), with accountability for the service’s response to each concern at its centre; this responds to neutralisation by making the decision not to act visible. The remaining proposals are grouped by the mechanism they counter. Community-generated evidence, produced by people and in spaces women trust, and qualitative routes, which ask women directly about racism and record their own accounts, counter discounting. Independent observation and ethnicity-disaggregated service delivery data counter neutralisation; the first moves the judgement of whether evidence is sufficient outside the service, and the second addresses deflection. Conditions for action counter containment, while conditions for safety allow evidence to be raised at all.

**Figure 2.**
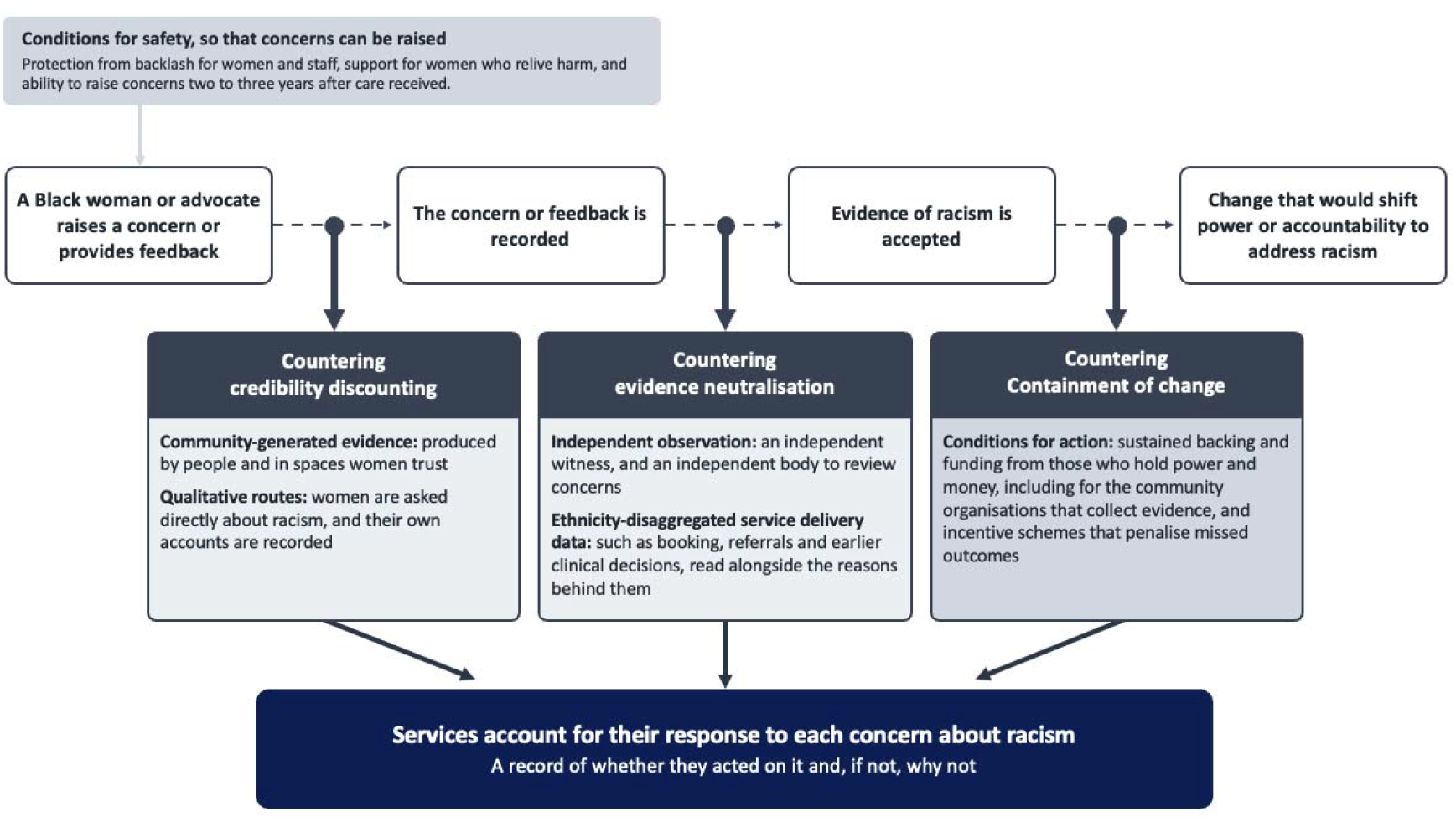
Black mothers’ and advocates’ proposals for recording evidence of racism for action

#### 3.3.1. Accounting for the response to racism

Participants argued that services should account for what they did with evidence of racism; one mother proposed the most concrete form:

> *‘I would say it should be recorded every time a woman asks for something or raises a concern… And what also needs to be recorded on the professional side is if they acted on that information and if they didn’t act on it, why didn’t they act on it?’* – P9 (mother)

Another mother in the same group argued that staff who had to record a decision not to act would be less willing to make that decision. Advocates in both groups called for feedback to be taken back to services and for inspection of whether organisations had acted on it; one proposed that regulators ask for this evidence and count it towards a service’s rating.

#### 3.3.2. Producing and reading the evidence

The remaining proposals describe who should produce, witness and read this record. Participants placed most weight on evidence produced outside the service, by communities or independent observers, because they did not expect services to recognise racism in their own records. One advocate explained this in terms of whose accounts are believed:

> *‘People will tell you. And when they tell you, we need to believe them… we’ve got the data of how many people are dying, but we haven’t got the data of the near misses in that national data… They [services] need to listen, to believe us. We’re not making it up, we’re not trying to do a hype. It is what it is.’* – P12 (advocate)

Qualitative routes concerned what is asked and recorded. Advocates argued that services were ‘not capturing the stories, the experiences’ (P13, advocate), and that racism would go unrecorded unless women were asked about it directly. Mothers wanted a choice of routes, including questionnaires for those who ‘might not want to talk face to face’ (P1, mother). Yet an account on its own remains open to being discounted as anecdotal, which may explain why participants paired it with independent observation and service data.

Independent observation addressed this by changing who witnesses care. Participants in several groups proposed an independent witness who could accompany women and see whether they were treated differently, likening this to mystery shopping, and an independent body to review concerns. This approach, however, relies on a third party being believed rather than changing who is believed.

Participants also wanted service data broken down by ethnicity, such as the timing of antenatal booking, referrals and access to services. Mothers and advocates, however, wanted different things from these data. Mothers saw numbers as part of a written record that would protect them from being disbelieved: ‘always statistics, emails, letters, everything that you can seize in black and white, because then it can’t be my word against yours’ (P9, mother). Two advocates were wary of numbers replacing women’s accounts, although both said they used ethnicity-disaggregated measures themselves. Advocates also wanted data on earlier decisions, such as offers of induction, since caesarean rates are ‘the last stop’ and earlier intervention ‘gets explained away that that’s normal’. Hospitals held these numbers, ‘but what they don’t have is the why, as in the honest why of it’ (P15, advocate). Participants therefore tended to treat numbers as evidence of racism when read alongside the reasons behind them and by someone outside the service.

#### 3.3.3. Conditions for safety and for action

Participants said recording alone would not be enough and described conditions for safety, so that evidence could be raised, and for action, so that it would be used. The first condition for safety was protection from backlash, as mothers described holding evidence back for fear it would affect their care. One proposed keeping a feedback log during pregnancy to hand in at the end:

> *‘…If someone gives you negative feedback, you will feel a way, if you were a midwife, reading something that your patient is basically saying that they don’t feel like you’re meeting their needs, midway through the journey. So, I guess it’s something that we, as the parents, or mums, could keep with us, and then hand it in, maybe, at the end?’* – P2 (mother)

Advocates described the same fear among staff, as midwives who saw women treated unfairly could not escalate it without risk to themselves. Safety also meant support, as a traumatised mother should not be asked to relive her experience ‘without her having that additional support to hold her’ (P15, advocate). It also meant time, as advocates proposed that monitoring continue for two to three years after birth because ‘it takes time to acknowledge that harm was experienced’ (P11, advocate).

Conditions for action concerned who backs the evidence and who pays for it. Advocates argued that evidence would lead to change only with sustained backing from those who hold power and money:

> *‘[…] you have to have a mandate and you have to have support higher up the ladder, like in commissioning services and ICBs (integrated care boards)… without that backing from high up, you won’t get that far, or a lot of the work you would do would be noticed for a little while, it becomes flavour of the month or flavour of the week, and then what happens after that?’* – P10 (advocate)

Advocates also called for sustained funding for the community organisations that collect evidence, often staffed by underfunded or part-time advocates, and for incentive schemes that penalise missed outcomes as well as rewarding targets met. Their remedies for containment were therefore redistributive, requiring a transfer of money and authority that better evidence alone would not achieve.

## 4. DISCUSSION

Drawing on the accounts of Black mothers and community advocates, we identified three mechanisms, credibility discounting, neutralisation and containment, that may help explain why evidence of racism in maternity care often does not lead to action. Unlike national investigations, which cover maternity care for all service user groups [8,10], and research drawing largely on staff accounts [12,13,54,55], this study offers an explanation of how evidence of racism in particular is handled, from the perspectives of Black mothers and community advocates.

The credibility discounting participants described is consistent with UK Black-led and community inquiries into English NHS maternity services [5,7] and with evidence from other countries such as Canada, Brazil and the United States [58–61]. Women in these studies also held back concerns; in one UK survey, around half of Black women who experienced poor care did not complain formally, often believing nothing would change or fearing worse care [7]. This suggests that complaint numbers understate racism before any concern reaches a service. Advocacy sometimes prompted action, but only by substituting the advocate’s credibility for the woman’s. This offsets discounting in individual cases without correcting it and risks individualising responsibility for navigating inequitable systems [18].

Our findings also bring together research on epistemic injustice, which shows how patients’ testimony is given less credibility than it deserves [45,56], and on audit culture, which shows how processes designed to demonstrate accountability can come to stand in for it [50–52]. Audit culture research has focused mainly on how organisations and professionals perform accountability, whereas our findings suggest that, for evidence of racism, what enters the audit also depends on whose testimony is believed. A record thinned by discounting is easier to absorb or deflect. In Jones’s terms, interpersonal racism thins the record on which institutional racism then operates, so the levels compound [39]. Dashboards now report gaps between ethnic groups within each service [9], but where inequalities run through the whole system, a service whose gap matches the national pattern can still show it is not an outlier. Linking incentives to such indicators also risks the target distortion participants described [49]. Outcome data also record harm only after it occurs, whereas women’s own accounts may be the closest equivalent to near-miss reporting.

Failure to implement recommendations has been attributed to their volume, overlap and lack of a clear owner [8], and to constrained resources and overlapping demands for data [57], so that reporting can consume the capacity it is meant to direct [52]. These explanations treat inaction mainly as a question of capacity. Containment suggests that, for racism, inaction may also reflect who controls resources. Ray’s theory of racialised organisations suggests that ordinary rules and routines can reproduce racism without anyone intending harm [46]. Our findings are consistent with this account, as services appeared to give Black women’s concerns less weight, keep complaints processes separate from practice and leave control of resources unchanged.

Our findings suggest why remedies for defensive culture may not address racism. Efforts to address defensive culture in the NHS have focused on encouraging staff to speak up and services to be open when things go wrong [12,13,54], whereas participants’ conditions for safety suggest that women who raise concerns need similar protection from backlash. For evidence of racism, the problem may lie earlier, as discounted concerns may not reach the processes these efforts rely on. Where services respond most to those expected to complain, greater openness may still leave Black women’s concerns unaddressed. Framing racism as a patient safety issue may have similar limits, as it places racism within governance that relies on internal reporting. This governance sits alongside clinical negligence processes, which the national investigation found can create incentives to deny problems and challenge families’ accounts [8]. These responses work within existing services, leaving unchanged who judges whether evidence is sufficient to act on.

Harm from staff who shared women’s racial identity adds an intersectional dimension to these mechanisms, as the hierarchy mothers described operated within racial groups as well as between them [25,53], which monitoring by broad ethnic category would miss. Advocates’ accounts of Black midwives being dismissed in the same way as the women they cared for suggest that Black staff are themselves subject to the racialised organisations they work in [46]. Participants who reviewed the findings linked this harm to the organisation, as staff who face racism at work may treat women as they are treated. A more diverse workforce may not, on its own, change how Black women and families are cared for.

Participants’ proposals respond to limits in existing measures. Patient-reported experience measures, including measures of obstetric racism [16], and the experience data that NHS maternity boards review [62] may undercount racism where Black women hold back concerns or are not believed, and they record what women experienced rather than what services did in response. The proposals address these limits only in part. A record of the response to each concern would still be produced within services that discount Black women’s accounts, and like other accountability measures it could come to stand in for action [14,50–52]. Independent observation substitutes a credible witness for the woman, as advocacy does, which leaves her own testimony no more likely to be believed [45]. These proposals also leave unchanged who controls the resources to act.

Participants’ conditions for action are consistent with evidence that stable funding and power-sharing strengthen community advocates’ capacity to challenge inequities, although advocacy alone cannot resolve systemic disparities [18]. Short-term or externally controlled funding can compromise the independence of community organisations [18], as can contracting through procurement and performance management [63]. Funding would therefore need to be long-term and leave these organisations free to challenge the services they work with. It should also avoid adding to the unpaid, emotionally taxing work of evidencing racism that already falls on Black women and advocates [64,65].

### 4.1. Implications for policy, practice, and future research

Our findings suggest that more measurement of racism alone will not change care. To counter credibility discounting, services could ask women directly about racism and record their accounts alongside complaints and survey scores, as these accounts may be the only record of near misses. They could also invite women to raise concerns through community organisations, and for several years after their care has ended, as recognising harm takes time. To counter neutralisation, board reports on safety and experience data [8,62] could record the response to each concern about racism and the reason for any decision not to act. Ethnicity data could cover earlier decisions such as referrals, with the reasons for them, using categories that reflect the diversity of women served [8]. Some participants proposed that regulators such as the Care Quality Commission review this record. Regulators are not neutral readers, however; the national investigation heard of families ignored in services rated ‘good’ and asked the Nursing and Midwifery Council to check staff fitness to practise referrals for racial bias [8]. Community organisations working with Black families should therefore review this reporting alongside regulators and share decisions on how services respond. To counter containment, commissioners and integrated care boards could move funding for gathering evidence of racism to community organisations that support Black families, on a long-term basis that protects their independence, while services remain accountable for acting on what they find. Services should also tell those who raised concerns what was done, as the belief that nothing will change leads women to hold concerns back [7].

Because we did not compare how services handled evidence of racism with other service failures, future studies could test whether these mechanisms are specific to evidence of racism, for example by comparing complaint outcomes by ethnicity and type of concern. Research with maternity staff and boards could show how the mechanisms operate inside services. Research with migrant Black women and Black staff could test whether the hierarchy described in one group operates through the suggested mechanisms. The participants’ framework could be developed into a practical tool and evaluated, including the burden it places on women and advocates. Comparing health systems that fund and govern community advocacy differently could show whether moving funding to community organisations changes how evidence of racism is used.

### 4.2. Strengths and limitations

Lived experience co-researchers shaped the design, facilitation and interpretation, sharing the judgement between accounts that in critical realist research often rests with the researcher [30]. Co-facilitation by Black women may have made it easier for participants to describe anti-Black racism with a non-Black researcher present [26,27]. Including mothers and advocates combined women’s accounts of their own care with advocates’ accounts of how services handle concerns across many families. The analysis aimed to explain rather than describe, and each mechanism was tested against rival explanations.

The mechanisms are proposed explanations, and we do not claim that they are exhaustive or that they operate in the same way in other settings. The study relies on four focus groups of mixed size in one city. It did not include staff or service perspectives, so the containing and neutralising mechanisms rest partly on advocates’ and mothers’ views of how services are run. Data collection coincided with the publication of reports from two independent investigations into NHS maternity care [8,10], which may have shaped advocates’ accounts. We also did not compare how services handled evidence of racism with other service failures, so the claim that racism faces barriers beyond a general defensive culture rests on participants’ accounts. Some participants knew the lived experience co-researchers or the first author, which may have made it more comfortable to name racism as an explanation and describe experiences that might otherwise have gone unreported. We addressed this through reflexive memos and shared interpretation with the lived experience co-researchers.

Finally, London has more community organisations working with Black families than most of England, so participants may have been better able to name racism than families elsewhere. Several mechanisms, however, operate through national structures such as complaints processes and incentive schemes, and we argue transferability through these mechanisms rather than the representativeness of the sample [36].

## 5. CONCLUSION

Evidence of racism in maternity care is increasingly collected but seldom leads to action. Black mothers’ and community advocates’ accounts suggest why. Black women’s concerns may be discounted before they are recorded, recorded evidence absorbed or deflected through the processes services use to demonstrate performance, and change that would shift power or accountability withheld. More evidence alone is therefore unlikely to change care, as participants asked who decides when evidence is enough. They proposed that services record what they do with each concern, that this record be reviewed independently and that those who raise concerns be protected, making visible a decision not to act. Change is more likely where services also shift power and money to community organisations working with Black families, while remaining accountable for acting on evidence of racism.

## Data Availability

The focus group transcripts are not publicly available because they contain detailed accounts of individual care that could identify participants, and participants did not consent to public data sharing.

## 6. LIST OF ABBREVIATIONS

CIC: community interest company
GRIPP2: Guidance for Reporting Involvement of Patients and the Public, version 2
ICB: integrated care board
NHS: National Health Service
SRQR: Standards for Reporting Qualitative Research

## 7. ADDITIONAL FILES

**Additional file 1:** Reporting of co-production using the GRIPP2 short form (GRIPP2-SF).

**Additional file 2:** Study reporting as per Standards for Reporting Qualitative Research (SRQR).

**Additional file 3:** Final version of the semi-structured topic guide used in the focus groups.

## 8. DECLARATIONS

## Ethics approval and consent to participate

The study received a favourable opinion from the Imperial College Research Ethics Committee (ref. 8208475) and was conducted in accordance with the Declaration of Helsinki. All participants gave consent before taking part.

## Consent for publication

Not applicable. The manuscript contains no identifiable individual data; quotations are pseudonymised and attributed by participant number and role only.

## Availability of data and materials

The focus group transcripts are not publicly available because they contain detailed accounts of individual care that could identify participants, and participants did not consent to public data sharing. The topic guide is provided in Additional file 3.

## Authors’ contributions

Study design and conceptualisation were led by SA and the lived experience co-researchers (SI, RNM, PF, CW and GS) and with support from PA, SD and HW. The focus group topic guide was co-produced by SA, SI, RNM, PF, CW and GS. Study recruitment was conducted by SA and RW, with SI, RNM, PF, CW and GS sharing the study through their networks. The focus groups were co-facilitated by SI, RNM, PF and CW, with support from SA. Coding was conducted by SA, RW, SI, RNM, PF, CW and GS, and SA coded the full transcripts. SA led the analysis and interpretation, and all co-authors reviewed and challenged interpretations. The manuscript was written by SA with critical review and edits from all authors. All authors read and approved the final manuscript.

## Acknowledgements

We thank all the participants who took part in this study for trusting us with their experiences, and the community organisations and networks that shared the study on our behalf.

