## Additional File 1 for "*‘When will it ever be enough?’* Black mothers’ and community advocates’ perspectives on why evidence of racism in English maternity care does not lead to action: a co-produced focus group study"

**ADDITIONAL FILE 1. REPORTING OF CO-PRODUCTION USING THE GRIPP2 SHORT FORM (GRIPP2-SF)**

The five lived experience co-researchers were members of the research team and are co-authors of this paper, a role that extends beyond patient and public involvement (PPI) as GRIPP2 defines it. We use the GRIPP2 short form [1] to report their contribution.

| **Section and topic** | **GRIPP2-SF item** | **Reported in this study** |
| --- | --- | --- |
| **1. Aim** | *Report the aim of PPI in the study.* | To share decisions with Black, African, Caribbean and mixed-Black community members across the study, so that the questions asked, the way they were asked and the reading of the findings reflected the priorities of people affected by racism in maternity care. |
| **2. Methods** | *Provide a clear description of the methods by which patients and the public were involved.* | Five lived experience co-researchers, who had used NHS maternity care or supported Black families as advocates, have worked with SA since March 2024, starting with the co-production prioritisation workshops from which this study arose, in which Black families, NHS professionals and public health teams in North West London identified community maternity advocates as a priority [2]. They were paid £27.50 an hour under the NIHR payment policy, trained in research ethics, trauma-informed practice and facilitation, and met with SA every fortnight. They contributed to the study design, piloted the topic guide, shared recruitment through their networks, co-facilitated the focus groups, took part in coding workshops and interpretation meetings, and are study co-authors; three also co-presented with SA at the findings review sessions with participants. |
| **3. Results** | *Report the outcomes of PPI in the study, including both positive and negative outcomes.* | Each focus group was co-facilitated by two lived experience co-researchers, which contributed to rich discussion in which participants readily described their own experiences. The co-researchers helped revise the topic guide for clarity after the first two focus groups. They also changed the study in specific ways: the first research question was reworded into plain language; the harm from staff who share women’s racial identity was retained in the findings at their request, where the first author had questioned its relevance, and reframed around diaspora and time in the UK; collective spaces were added to the qualitative routes; the neutralising mechanism was extended after co-researchers proposed that staff and services may be excused because of a visible record of anti-racism work, such as completed training; and community-generated evidence was read as a double bind, which informed the Discussion of the burden and backlash it can place on women and advocates. Negative outcomes: a lived experience facilitator introduced a label for harm from Black staff that participants then adopted in one session; we therefore did not use the term and grounded this finding in participants’ own accounts. |
| **4. Discussion and conclusions** | *Comment on the extent to which PPI influenced the study overall. Describe positive and negative effects.* | Co-production shaped the research questions, the facilitation and the interpretation of findings, including the decision to retain the finding on harm from staff who share women’s racial identity, which informed the Discussion point that a more diverse workforce may not on its own protect Black women, and the extension of the neutralising mechanism to include a visible record of anti-racism work. It had less influence on the academic theoretical framing used, including the choice of critical realism and the theories used in retroduction, which the first author introduced. Co-facilitation by community members may have made it easier for participants to share their own experiences, which many did although the questions did not ask them to do so directly. |
| **5. Reflections and critical perspective** | *Comment critically on the study, reflecting on the things that went well and those that did not, so others can learn from this study.* | **What went well:** A relationship built over two years, with fair payment and training, made co-facilitation easier, and participants often directed their accounts to the co-researchers. Debriefs gave co-researchers a route to challenge the design and also to provide peer-to-peer emotional support during data collection and analysis. Some of the lived experience co-researchers recognised audit culture from their own experience, suggesting the theory named something familiar rather than imposed.  **What could be improved:** Review time was limited and attendance uneven because of a clash with summer holidays and project timelines, so the first author met individually with those who missed sessions; introducing the critical realist paradigm to the lived experience co-researchers required additional training during study development. Reviewing the findings was emotionally demanding for co-researchers, whose own experiences informed findings only where participant data supported them. Others should plan sufficient review time and individual follow-up, present findings in plain language, and support co-researchers emotionally during analysis. |

Reference numbers refer to the reference list of the main manuscript.

NIHR, National Institute for Health and Care Research; PPI, patient and public involvement.

***References:***

*1. Staniszewska S, Brett J, Simera I, Seers K, Mockford C, Goodlad S, et al. GRIPP2 reporting checklists: tools to improve reporting of patient and public involvement in research. Res Involv Engagem. 2017;3:13. https://doi.org/10.1186/s40900-017-0062-2*

*2. Aryasinghe S, Averill P, Waithe C, Ibuanokpe S, Newby-Mayers R, Lakhdar N, et al. Improving the maternity experience for Black, African, Caribbean and mixed-Black families in an integrated care system: a multigroup community and interprofessional co-production prioritisation exercise using nominal group technique. BMJ Qual Saf. 2025;34:305–16. https://doi.org/10.1136/bmjqs-2024-017848*
