## Additional File 2 for "*‘When will it ever be enough?’* Black mothers’ and community advocates’ perspectives on why evidence of racism in English maternity care does not lead to action: a co-produced focus group study"

**ADDITIONAL FILE 2. STUDY REPORTING AS PER STANDARDS FOR REPORTING QUALITATIVE RESEARCH (SRQR).**

*Study reporting against the Standards for Reporting Qualitative Research (SRQR) [1]*

| **No.** | **SRQR item** | **SRQR guidance** | **How it is reported in this manuscript** | **Section (line numbers)** |
| --- | --- | --- | --- | --- |
| **Title and abstract** | | | | |
| 1 | Title | Concise description of the nature and topic of the study. Identifying the study as qualitative or indicating the approach (e.g., ethnography, grounded theory) or data collection methods (e.g., interview, focus group) is recommended | The title names the topic, the participants (Black mothers and community advocates), the co-produced design and the focus group method. | Title page |
| 2 | Abstract | Summary of key elements of the study using the abstract format of the intended publication; typically includes background, purpose, methods, results, and conclusions | Structured abstract with Background (including the aim), Methods, Results and Conclusions, and ten keywords. | Abstract (lines 1–35) |
| **Introduction** | | | | |
| 3 | Problem formulation | Description and significance of the problem/phenomenon studied; review of relevant theory and empirical work; problem statement | Racism in UK maternity care and the higher maternal mortality of Black women; reliance on complaints, patient experience surveys and the Maternity and Neonatal Equalities Dashboard; the national investigation’s framing of racism as a safety issue; and existing explanations for why evidence goes unused, which draw mainly on staff accounts. | 1. Background (lines 38–75) |
| 4 | Purpose or research question | Purpose of the study and specific objectives or questions | Two research questions, from the perspective of Black mothers and community advocates, on what appears to stop the monitoring of racism in maternity care from leading to action, and how evidence of racism should be recorded so that it does. | 1. Background, final paragraph (lines 68–75) |
| **Methods** | | | | |
| 5 | Qualitative approach and research paradigm | Qualitative approach (e.g., ethnography, grounded theory, case study, phenomenology, narrative research) and guiding theory if appropriate; identifying the research paradigm (e.g., postpositivist, constructivist/ interpretivist) is also recommended; rationale** | A co-produced qualitative focus group study underpinned by critical realism (research paradigm) and Black feminist thought (guiding theory). The rationale for combining them, the tension between the authority of lived experience and mechanisms that may go unrecognised, and how shared interpretation with lived experience co-researchers addressed this are described. Analysis used Fryer’s critical realist thematic analysis. | 2.1 Study design and theoretical approach (lines 77–104); 2.7 Coding and data analysis (lines 165–191) |
| 6 | Researcher characteristics and reflexivity | Researchers’ characteristics that may influence the research, including personal attributes, qualifications/experience, relationship with participants, assumptions, and/or presuppositions; potential or actual interaction between researchers’ characteristics and the research questions, approach, methods, results, and/or transferability | The first author’s ethnicity, role in a national anti-racism organisation in health and the risk this posed to interpretation, and her lack of shared experience of anti-Black racism and NHS maternity care. The roles of the five Black lived experience co-researchers, RW and the supervisors in developing and challenging interpretations. Prior relationships with some participants are reported as a limitation. | 2.2 Our positionality and reflexivity (lines 105–120); 4.2 Strengths and limitations (lines 611–615) |
| 7 | Context | Setting/site and salient contextual factors; rationale** | London, chosen for its higher proportion of Black, African, Caribbean or mixed-Black residents than elsewhere in England, so that participants could describe care across several maternity services. Groups were held at a south London community venue and on Zoom. The London context and the timing of two national maternity reviews are discussed as limitations. | 2.3 Study setting (lines 121–124); 2.6 Data collection and processing (lines 157–158); 4.2 Strengths and limitations (lines 607–609, 616–617) |
| 8 | Sampling strategy | How and why research participants, documents, or events were selected; criteria for deciding when no further sampling was necessary (e.g., sampling saturation); rationale** | Purposive sampling of Black, African, Caribbean or mixed-Black service users aged 18 or over who had used London NHS maternity services in the past five years, and of advocates of any ethnicity supporting Black families in London. Recruitment routes, screening (including for fraudulent participation) and the rationale for separate groups are described. Sample adequacy was judged by information power rather than saturation. | 2.5 Participants and recruitment (lines 131–148) |
| 9 | Ethical issues pertaining to human subjects | Documentation of approval by an appropriate ethics review board and participant consent, or explanation for lack thereof; other confidentiality and data security issues | Imperial College Research Ethics Committee approval (ref. 8208475); signed electronic consent; withdrawal up to 14 days after the session; limits to confidentiality; support before, during and after sessions; trauma-informed facilitation and scenario-based questions; pseudonymised recordings on encrypted servers. | 2.4 Ethics approval (lines 125–130); 2.6 Data collection and processing (lines 149–164); 8. Declarations (lines 647–651) |
| 10 | Data collection methods | Types of data collected; details of data collection procedures including (as appropriate) start and stop dates of data collection and analysis, iterative process, triangulation of sources/methods, and modification of procedures in response to evolving study findings; rationale** | Four semi-structured focus groups (two with mothers, two with advocates) held between 19 June and 3 July 2026, co-facilitated by two lived experience co-researchers with SA and RW, with debriefs after each. Data were analysed from July to October 2026. Focus groups were chosen because dialogue among those who share identities can make racism easier to discuss and recognise; including both groups set women’s accounts of their own care alongside advocates’ accounts across many families. | 2.1 Study design and theoretical approach (lines 90–93); 2.6 Data collection and processing (lines 149–164); 2.7 Coding and data analysis (lines 165–191); 4.2 Strengths and limitations (lines 600–602) |
| 11 | Data collection instruments and technologies | Description of instruments (e.g., interview guides, questionnaires) and devices (e.g., audio recorders) used for data collection; if/how the instrument(s) changed over the course of the study | A semi-structured topic guide co-produced with the lived experience co-researchers and revised for clarity after the first two groups (Additional file 3); an online screening questionnaire; a dictaphone for in-person groups and Zoom for online groups. | 2.6 Data collection and processing (lines 152–158); 2.5 Participants and recruitment (lines 137–138); 7. Additional files (line 644) |
| 12 | Units of study | Number and relevant characteristics of participants, documents, or events included in the study; level of participation (could be reported in results) | Sixteen participants (nine mothers, seven advocates) in four groups of two to six; two groups were smaller than planned because of drop-outs on the day. Characteristics are given in Table 2 and group composition in Table 3. | 3.1 Participant characteristics; Tables 2 and 3 (lines 196–206) |
| 13 | Data processing | Methods for processing data prior to and during analysis, including transcription, data entry, data management and security, verification of data integrity, data coding, and anonymization/de-identification of excerpts | Pseudonymised recordings on encrypted servers, professionally transcribed after the withdrawal period; transcripts checked against recordings and de-identified by SA; coding in Microsoft Word and NVivo v.15; quotations attributed by participant number and role (mother or advocate). | 2.6 Data collection and processing (lines 162–164); Table 1, Step 2 (line 192); 8. Declarations (lines 650–651) |
| 14 | Data analysis | Process by which inferences, themes, etc., were identified and developed, including the researchers involved in data analysis; usually references a specific paradigm or approach; rationale** | Fryer’s five-step critical realist thematic analysis (Table 1), chosen because it produces causal explanations rather than descriptions of experience. Codes were consolidated through abduction and themes developed through retroduction, starting from participants’ own causal explanations and testing them against the events described and existing theory. SA, RW and the lived experience co-researchers coded together; SA coded all transcripts and led the interpretation. | 2.7 Coding and data analysis (lines 165–191); Table 1 (line 192) |
| 15 | Techniques to enhance trustworthiness | Techniques to enhance trustworthiness and credibility of data analysis (e.g., member checking, audit trail, triangulation); rationale** | Each mechanism was tested against rival explanations and all transcripts were searched for cases in which the expected outcome did not occur. Codes were reviewed for interpretative validity and themes for theoretical validity. Joint interpretation with the lived experience co-researchers, reflexive memos and challenge from co-authors were used in place of inter-coder agreement. As set out in the study protocol, key findings were shared with participants for review; SA, SI, PF and CW presented them at review sessions with interested participants, which refined the mechanisms. | 2.7 Coding and data analysis (lines 180–188); Table 1, Step 4 (line 192); 2.2 Our positionality and reflexivity (lines 105–120); 2.1 Study design and theoretical approach (lines 98–103) |
| **Results/findings** | | | | |
| 16 | Synthesis and interpretation | Main findings (e.g., interpretations, inferences, and themes); might include development of a theory or model, or integration with prior research or theory | Three proposed mechanisms (credibility discounting, neutralisation through audit culture, and containment of change that would shift power or accountability), with harm from staff who share a racial identity as an outcome (Figure 1). Participants’ proposals are synthesised in Figure 2. Each mechanism is presented with participants’ explanations, rival explanations and its theoretical grounding. | 3.2 and 3.3 Results (lines 207–495); Figures 1 and 2 (lines 215–216, 425) |
| 17 | Links to empirical data | Evidence (e.g., quotes, field notes, text excerpts, photographs) to substantiate analytic findings | Quotations and group exchanges, labelled by participant number and role, support each mechanism and proposal; disagreements and a case in which discounting was countered are reported. | 3.2 and 3.3 Results (lines 218–495) |
| **Discussion** | | | | |
| 18 | Integration with prior work, implications, transferability, and contribution(s) to the field | Short summary of main findings; explanation of how findings and conclusions connect to, support, elaborate on, or challenge conclusions of earlier scholarship; discussion of scope of application/generalizability; identification of unique contribution(s) to scholarship in a discipline or field | A summary of findings and contribution; comparison with research on epistemic injustice, audit culture, implementation of recommendations, defensive culture, racialised organisations, patient-reported experience measures and community advocacy; how far participants’ proposals address the mechanisms; and implications for policy, practice and research. Transferability is argued through the national structures the mechanisms operate through rather than the representativeness of the sample. | 4. Discussion (lines 496–567); 4.1 Implications for policy, practice, and future research (lines 568–595); 4.2 Strengths and limitations (lines 616–620) |
| 19 | Limitations | Trustworthiness and limitations of findings | Mechanisms are proposed explanations and not exhaustive; a small number of mixed-size groups in one city, with London’s community organisations possibly helping participants to name racism; no staff or service perspectives; data collection alongside national reviews; no comparison with other service failures; prior relationships between some participants and the research team. | 4.2 Strengths and limitations (lines 596–620) |
| **Other** | | | | |
| 20 | Conflicts of interest | Potential sources of influence or perceived influence on study conduct and conclusions; how these were managed | SA’s employment by the NHS Race and Health Observatory, which did not fund or oversee the study but shares a strategic focus with its subject matter, is declared; how this may have influenced interpretation is addressed in the positionality statement. | 2.2 Our positionality and reflexivity (lines 105–120); 8. Declarations, Competing interests (lines 655–656) |
| 21 | Funding | Sources of funding and other support; role of funders in data collection, interpretation, and reporting | Author support from the NIHR Imperial Biomedical Research Centre, the Collaboration Kickstarter Legacy Fund (Imperial College London), a Better Health & Care Hub Postdoctoral Fellowship (King’s College London), the Wellcome Trust and the Medical Research Council. Funders had no role in design, data collection, analysis, interpretation or writing. | 8. Declarations, Funding (lines 657–664) |

**The rationale should briefly discuss the justification for choosing that theory, approach, method, or technique rather than other options available, the assumptions and limitations implicit in those choices, and how those choices influence study conclusions and transferability. As appropriate, the rationale for several items might be discussed together.
