## Additional File 3 for "*‘When will it ever be enough?’* Black mothers’ and community advocates’ perspectives on why evidence of racism in English maternity care does not lead to action: a co-produced focus group study"

**ADDITIONAL FILE 3. FINAL FOCUS GROUP TOPIC GUIDE**

*Note: wording for advocates’ groups is shown where it differed. Facilitators adapted prompts to the flow of discussion.*

**Welcome and introduction (10 minutes)**

The facilitator introduces themselves as a lived experience partner and co-researcher on the project, and the first author as a PhD researcher at Imperial College London who will take notes and keep time. The facilitator explains that the project is about how community advocacy can help improve maternity care for Black, African, Caribbean and mixed-Black families. ***In person, the researcher points out fire exits, toilets and the quiet space.***

*For advocates: “We’ve asked you to join because you support Black communities during their maternity journey.”*

Today we want to hear your thoughts about

- What good and fair maternity care looks and feels like for Black families.
- How racism shows up in maternity care, from the perspective of Black families or advocates.
- What communities think should be recorded to hold maternity services accountable for racism.

This is a conversation. There are no right or wrong answers. Feel free to agree, disagree, or build on what someone else says.

**Ground rules**

- For online groups, raise your Zoom hand to speak and stay on mute.
- You can take a break, step out, or skip a question at any time. Your wellbeing comes first.
- Everyone’s perspective matters.
- You’re in control of what you share.
- Please keep anything shared here confidential.
- We know that some of you will have your own experiences of maternity care in addition to your maternity advocacy work. Please feel free to share as much or as little as you like about your own experiences.

**Does anyone want to add a ground rule?**

We’ll be recording today’s conversation so we can capture everything accurately. The recording is only for research, we won’t include names or anything that could identify you. **Is everyone happy for us to record?**

**Question 1 (20 minutes)**

**Can everyone say their name, and share what good and fair maternity care look and feel like for you? Share as much or as little as you’d like.**

- What makes care feel respectful? When does that matter most?
- Optional. Given the examples and experiences you’ve shared, what do you think needs to change so that every Black family receives good and fair maternity care?

*Transition. “Thank you, the experiences and examples you’ve shared, what good care looks like and where it falls short for Black families, are exactly the kind of information this study needs. Building on what you’ve shared just now, we want to understand what you think needs to be tracked or noticed for racism in maternity care to be taken seriously and acted on by the NHS. There are no right or wrong answers here; we’re really interested in what matters to you.”*

**Question 2 (20 minutes)**

**If an advocate was monitoring whether racism was happening in maternity services for Black families, what should they use as evidence?**

- What would they notice about how staff speak to or treat Black families?
- What would they notice about what happens when someone raises a concern?
- What would they notice about whether Black families are getting the care they need?

**Question 3 (15 minutes)**

**What evidence would you want to be collected that isn’t collected now?**

- How could this information be captured? At what stage (antenatal, labour and birth, postnatal)?
- How could this information be used to improve care?
- What evidence do you think is potentially overlooked or explained away?

**Question 4 (20 minutes)**

**If the evidence you’ve suggested was in place, how would you know it was being used meaningfully by the NHS?**

- What would need to be in place for that to happen?
- Why do you think this isn’t happening yet?

**Close (5 minutes)**

- Is there anything else you would like to say or add? (The researcher ends the recording.)
- Do you have any questions about today or the research?
